# Multiple indicators of gut dysbiosis predict all-cause and cause-specific mortality in solid organ transplant recipients

**DOI:** 10.1101/2023.10.28.23297709

**Authors:** J. Casper Swarte, Shuyan Zhang, Lianne M. Nieuwenhuis, Ranko Gacesa, Tim J. Knobbe, TransplantLines Investigators, Vincent E. de Meijer, Kevin Damman, Erik A.M. Verschuuren, C. Tji Gan, Jingyuan Fu, Alexandra Zhernakova, Hermie J.M. Harmsen, Hans Blokzijl, Stephan J.L. Bakker, Johannes R. Björk, Rinse K. Weersma

## Abstract

**Objective:** Gut microbiome composition is associated with multiple diseases, but relatively little is known about its relationship with long-term outcome measures. While gut dysbiosis has been linked to mortality risk in the general population, the relation with overall survival in specific diseases has not been extensively studied. In the current study, we present in-depth analyses regarding the relationship between gut dysbiosis and all-cause and cause-specific mortality in the setting of solid organ transplant recipients (SOTR).

**Design:** We analyzed 1,337 metagenomes derived from fecal samples of 766 kidney, 334 liver, 170 lung and 67 heart transplant recipients from the TransplantLines Biobank and Cohort; a prospective cohort study including extensive phenotype data with 6.5 years of follow up. To quantify gut dysbiosis, we included additional 8,208 metagenomic samples from a general population from the same geographical location. Multivariable Cox regression and a machine learning algorithm were used to analyze the association of indicators of gut dysbiosis and species abundances, with all-cause and cause-specific mortality.

**Results:** We identified two patterns representing overall microbiome community variation that were associated with both all-cause and cause specific mortality. Gut microbial distance to the average of the general population was associated with all-cause mortality and infection-, malignancy- and cardiovascular disease related mortality. Using multivariable Cox regression, we identified 23 species that were associated with all-cause mortality. By using a machine learning algorithm, we identified a log-ratio of 19 species predictive of all-cause mortality, all of which were also independently associated in the multivariable Cox-regression analysis.

**Conclusion:** Gut dysbiosis is consistently associated with mortality in SOTR. Our results support the observations that gut dysbiosis is predictive of long-term survival. Since our data do not provide causative evidence, further research needs to be done to see determine whether gut-microbiome targeting therapies might improve long term outcomes

**Summary box:** *Significance of this study:* What is already known on this subject?

- Current literature suggests that the gut microbiome signature might be associated with mortality risk in the general population.
- Higher diversity of gut microbiota is associated with lower mortality in allogeneic hematopoietic-cell transplantation recipients.
- Liver and kidney transplant recipients suffer from gut dysbiosis and an analysis with a relatively low number of events showed that dysbiosis is associated with mortality. What are the new findings?

- Across kidney, liver, heart and lung transplant recipients, we identified two overall microbial community variation patterns that are associated with all-cause mortality independent of the organ transplant and specifically to death from malignancy and infection.
- We find that multiple indicators of gut dysbiosis predict all-cause mortality and death by cardiovascular diseases, malignancy and infection.
- We find multiple microbial species associated with all-cause and cause-specific mortality. Using three different methods, we identify multiple bacterial species (shared between different analytical approaches) that are associated with an increased or decreased risk of mortality following solid organ transplantation.
- Using a machine learning algorithm, we identify a log-ratio of 19 bacterial species that was associated with all-cause mortality.

## Introduction

Gut dysbiosis, while not clearly defined, is a condition typically characterized by the growth of pathogens at the expense of commensal bacteria when compared to a healthy microbiome. A dysbiotic gut microbiome has been observed in many diseases, including inflammatory bowel disease, obesity, diabetes mellitus and cancer.^1–4^ Population-based studies report a large overlap in microbial associations to general health, suggesting a common dysbiotic signature in compromised health.^5–7^

Recent evidence suggests that such dysbiotic signatures are not only associated with a subject’s health status at time of sampling but also predictive of long-term survival. The gut microbiome, characterized by metagenomic sequencing of stool samples, was associated with mortality in a well-characterized population-based study of 7,211 adults with a follow-up of 15 years in Finland.^8^ This study found that members from the *Enterobacteriaceae* family were especially associated with an increased mortality risk.^8^ While studies linking gut dysbiosis with patient survival in specific disease populations are scarce, the relationship has been studied more extensively in allogeneic hematopoietic-cell transplantation where a lower alpha diversity is associated with increased mortality risk.^9,10^ We recently reported, in the setting of solid organ transplantation, that the extent of gut dysbiosis (in terms of the average distance from the general population) in both liver and kidney transplant recipients was associated with higher all-cause mortality risk.^11^ However, the number of events in this study was relatively small due to the limited follow-up time.

In the current study, we present an in-depth survival analysis of both all-cause and cause-specific mortality in a large population of solid organ transplant recipients (SOTR). In the current study, we have 3.7 times more samples and 2.7 times more events from liver, kidney, heart, and lung transplantation recipients compared with our previous analyses. This population of SOTR represents an appropriate model to study the relationship between gut dysbiosis and long-term survival, because the population SOTR is characterized by polypharmacy, multimorbidity and the prevalence of dysbiosis is high compared with the general population.^11,12^ Using this unique population of transplant recipients (n=1,337) and metagenomics samples from the general population (n=8,208), we analyzed the relationship between the gut microbiome and mortality. These findings are of interest for the transplantation community but also of interest for our general understanding of the gut microbiome and its relation to health.

## Results

### Characteristics of solid organ transplant recipients

In total, 1,337 solid organ transplant recipients who provided a fecal sample at variable time after transplantation, including 766 kidney (KTR), 334 liver (LTR), 170 lung (LuTR) and 67 heart (HTR) transplant recipients from the TransplantLines Biobank and Cohort study were included. The average age (± standard deviation, SD) of all recipients was 57 (± 13.0) years, 784 recipients (59%) were male and the average time since transplantation across all organ types was 7.6 (± 8.0) years (**Supplementary Figure 1A, 1B and 1C**). During the follow-up period with a minimum of 2.8 years and a maximum of 6.5 years, a total of 162 participants (88 KTR, 33 LTR, 35 LuTR and 6 HTR) died (Supplementary Table 1). Of these, 48 (28%) died due to infection related mortality, 38 (23%) due to cardiovascular related mortality, 38 (23%) due to malignancy related mortality, and 40 (25%) died from other causes (**Supplementary Table 1**).

In the first part of the analysis, we computed for each transplant recipient’s gut microbiome, indicators of gut dysbiosis: Shannon diversity index, the distance to the average microbiome composition of the general population, richness of antibiotic resistance genes (ARGs) and virulence factors (VFs). With multivariable Cox regression including age, sex, BMI and years since transplantation, we subsequently investigated the relationship between each of these indicators and recipient all-cause and cause-specific mortality. While we analyzed all-cause mortality for each transplant organ type separately, we only performed the cause-specific analysis on all SOTR pooled, because of smaller numbers of these events. In the second part of the analysis, we aimed at identifying microbial species that individually or jointly predict mortality. We investigated the relationship between transplant recipient mortality and each species’ CLR-transformed abundance, the quantile of each species in the general population. Lastly, we used a machine learning algorithm to identify which log-ratio of species best predict mortality.

### Overall community variation is associated with mortality risk

We first performed a Principal Component Analysis (PCA) on CLR-transformed species abundances. In all SOTR we observed a significant relationship between principal component (PC) 1 and increased all-cause mortality and decreased all-cause mortality and PC3 (PC1: HR=1.32, 95% CI=1.13-1.54, FDR=5.8×10^−4^; PC3: HR=0.80, 95% CI=0.68-0.93, FDR=4.0×10^−3^; **Figure 1A and 1B**). PC3 was also associated with a lower mortality for KTR and HTR (KTR: HR=0.73, 95% CI=0.59-0.89, FDR=2.2×10^−3^; HTR: HR=0.34, 95% CI=0.12-0.95, FDR=0.03; **Figure 1C**). In the cause-specific analysis, we found that PC1 was related to death from malignancy and infection (malignancy: HR=1.64, 95% CI=1.20-2.24, FDR=2.0 x 10^−3^; infection: HR=1.42, 95% CI=1.06-1.89, FDR=0.01; **Figure 1A, 1B and 1D**), and PC3 with a decreased risk of death from malignancy (HR=0.68, 95% CI=0.49-0.94, FDR=2.0 x 10^−2^). The five species that exhibited the largest positive loadings onto PC1 (and thus were associated with mortality) were *Ruminococcus gnavus, Clostridium clostridioforme, Clostridium symbiosum, Hungathella hathewayi*, and *Clostridium innocuum* (**Figure 1E**), and the five species that exhibited the largest positive loadings onto PC3 (and thus were inversely associated with mortality) *were Bifidobacterium adolescentis, Dorea longitena, Bifidobacterium longum, Collinsella aerofaciens* and *Eubacterium rectale* (**Figure 1F**).

**Figure 1.**
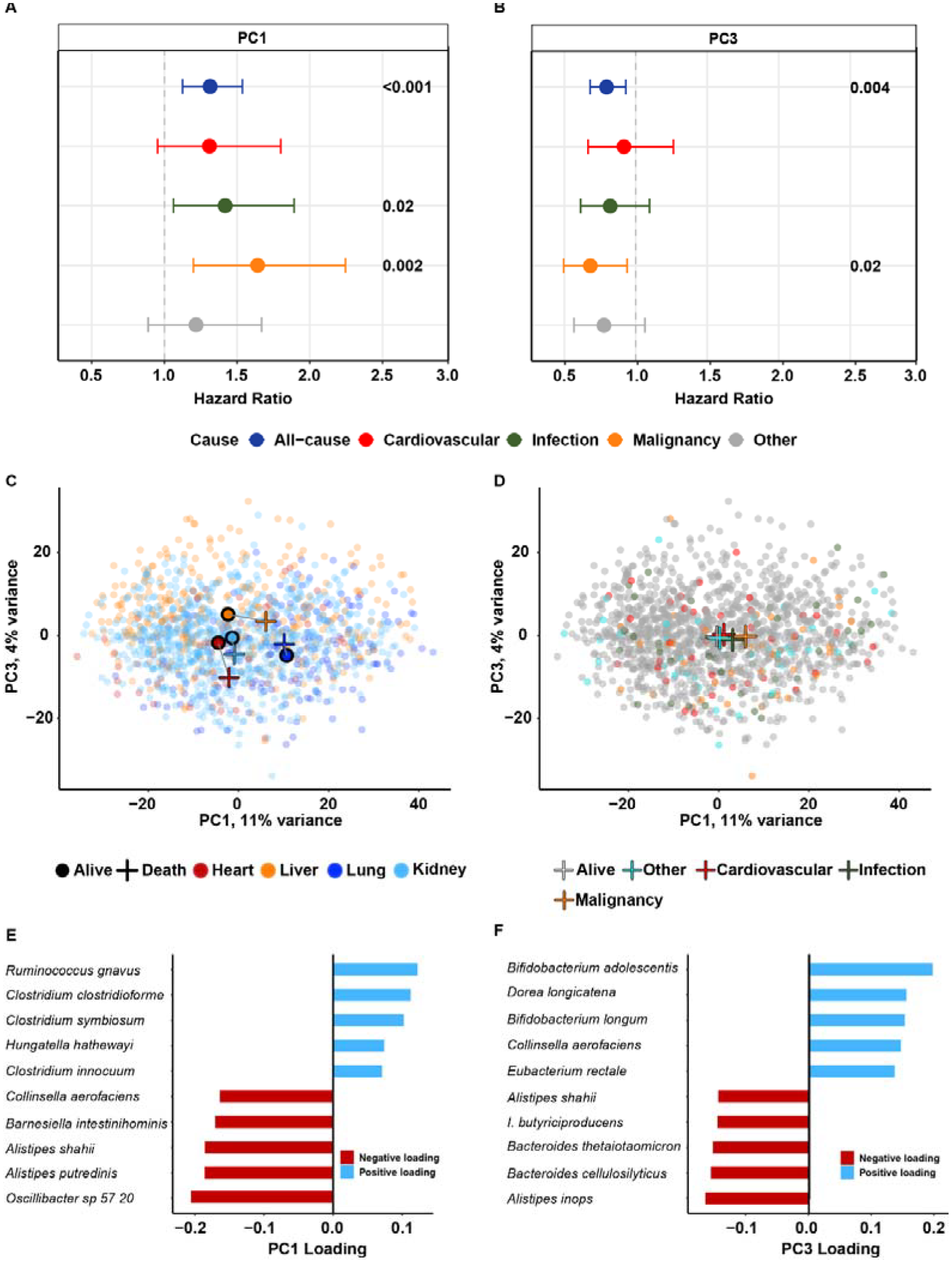
**(A and B)** Forest plot depicting results from Cox-regression analysis performed in principal component 1 and 3 for all-cause and cause-specific mortality. **(C)** Principal component analysis (PCA) colored by transplantation type. Circles and crosses represent centroids of SOTR that are alive and dead at the time of the follow-up, respectively. **(D)** PCA colored by cause of death showing the centroids (represented by crosses) of infection and malignancy related mortality are separated from cardiovascular and other causes of death in PC1. **(E and F)** Principal component loadings consisting of the 10 species with the largest positive and negative loadings for PC1 and PC3, thus were associated with mortality. I.: *Intestinimonas*.

### Multiple gut dysbiosis indicators are associated with all-cause and cause-specific mortality

In the cause-specific analyses performed in all SOTR, we found that the Shannon diversity index was related to death from malignancy (HR=0.71, 95% CI=0.54-0.93, FDR=0.01). We calculated how far each transplant recipient’s gut microbiome was from the average composition of the general population (Aitchison distance) and performed Cox-regression analyses. The distance to the general population was significantly associated with higher mortality risk in all SOTR (HR=1.29, 95% CI=1.11-1.51; FDR = 9.3 x 10^−4^; **Figure 2, Supplementary Figure 2**). In the cause-specific analyses, the distance to the general population was related to death from infection (HR=1.46, 95% CI=1.11-1.93, FDR=7.2×10^−3^), malignancy (HR=1.39, 95% CI=1.02-1.89, FDR=0.03) and cardiovascular disease (HR=1.36, 95% CI=1.01-1.87, FDR=0.04; **Figure 2**). Finally, we found that harboring a higher richness of antibiotic resistance genes (ARG) s and virulence factors (VFs) were associated with an increased all-cause mortality risk (ARGs: HR=1.27, 95% CI=1.09-1.47; FDR=2.2×10^−3^, **Figure 2**; VFs: HR=1.14, 95% CI=1.01-1.27; FDR=0.03; **Figure 2**) and with death from infection in the cause-specific analyses (ARGs: HR=1.45, 95% CI=1.10-1.90, FDR=8.0 x 10^−3^; VFs: HR=1.28, 95% CI=1.09-1.51, FDR=3.0 x 10^−3^; **Figure 2**). We did not observe a significant association between the Shannon diversity index and all-cause mortality when we analyzed recipients together and stratified by organ type (FDR>0.05, **Supplementary Table 2**). Overall, higher distance to the general population, increased in richness of ARG and VF, and decreased in Shannon diversity were linked to increased mortality.

**Figure 2.**
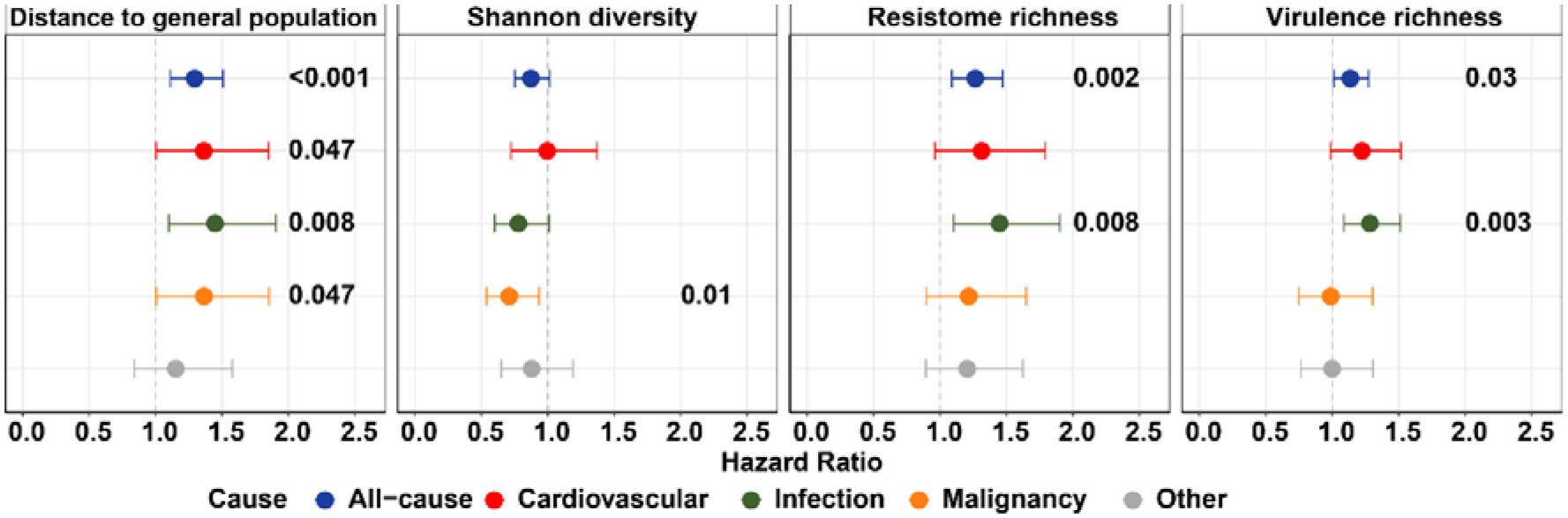
Alpha diversity metrics for all-cause and cause-specific mortality analysis. We calculated the Aitchison distance of each transplant recipient from the average composition of the general population and identified a relationship with all-cause and cause specific mortality. The Shannon diversity index was associated with malignancy related mortality. The richness of antibiotic resistance genes and virulence factors were both associated with all-cause and infection related mortality.

### Multiple species are associated with mortality

We identified a total of 23 (16%; FDR<0.10) species whose CLR-transformed abundance were associated with all-cause mortality using multivariate Cox-regression including age, sex, BMI and years since transplantation (**Supplementary Table 5**). When we analyzed all SOTR, we found four *Clostridium* species (*C. innocuum* [HR=1.35, 95% CI=1.18-1.54, FDR=0.001], *C. clostridioforme* [HR=1.34, 95% CI=1.16-1.54, FDR=0.003], *C. symbiosum* [HR=1.35, 95% CI=1.17-1.57, FDR=0.003] and *C. bolteae* [HR=1.32, 95% CI=1.157-1.52, FDR=0.004]) that were positively associated with all-cause mortality (**Figure 3A**; **Supplementary Table 5**) and death from infection in the cause-specific analyses (**Supplementary Table 6; Figure 3B**). Other species that were associated with an elevated mortality risk included *H. hathewayi* (HR=1.29, 95% CI=1.12-1.50, FDR=0.01), *Veillonella parvula* (HR=1.29, 95% CI=1.12-1.49, FDR=0.01) and *R. gnavus* (HR=1.26, 95% CI=1.09-1.46, FDR=0.03; **Figure 3A**; **Supplementary Table 5**). In the cause-specific analyses, we found that the abundance of *H. hathewayi* (HR=1.48, 95% CI=1.18-1.92, FDR=0.08) and *V. parvula* (HR=1.57, 95% CI=1.21-2.03, FDR=0.04) were related to death from infection (**Supplementary Table 6; Figure 3B**) and that the abundance of *R. gnavus* was related to death from malignancy (HR=1.83, 95% CI=1.34-2.49, FDR=0.04; **Supplementary Table 6; Figure 3B**). We also identified multiple species that were associated with a lower mortality risk when we analyzed all SOTR. For example, butyrate producers *Eubacterium hallii* (HR=0.75, 95% CI=0.63-0.89, FDR=0.01), *Firmicutes bacterium CAG 83* (HR=0.77, 95% CI=0.66-0.90, FDR=0.01), *Gemmiger formicilis* (HR=0.77, 95% CI=0.66-0.89, FDR=0.01) and *Faecalibacterium prausnitzii* (HR=0.83, 95% CI=0.73-0.95, FDR=0.05) were negatively associated with mortality (**Figure 3A**; **Supplementary Table 5**). Other commensals that were associated with a lower mortality risk included *Adlercreutzia equolifaciens* (HR=0.77, 95% CI=0.66-0.91, FDR=0.02), *Prevotella copri* (HR=0.77, 95% CI=0.65-0.91, FDR=0.03), *Asaccharobacter celatus* (HR=0.79, 95% CI= 0.67-0.93, FDR=0.04) and *D. longicatena* (HR=0.80, 95% CI=0.68-0.93, FDR=0.04).

**Figure 3.**
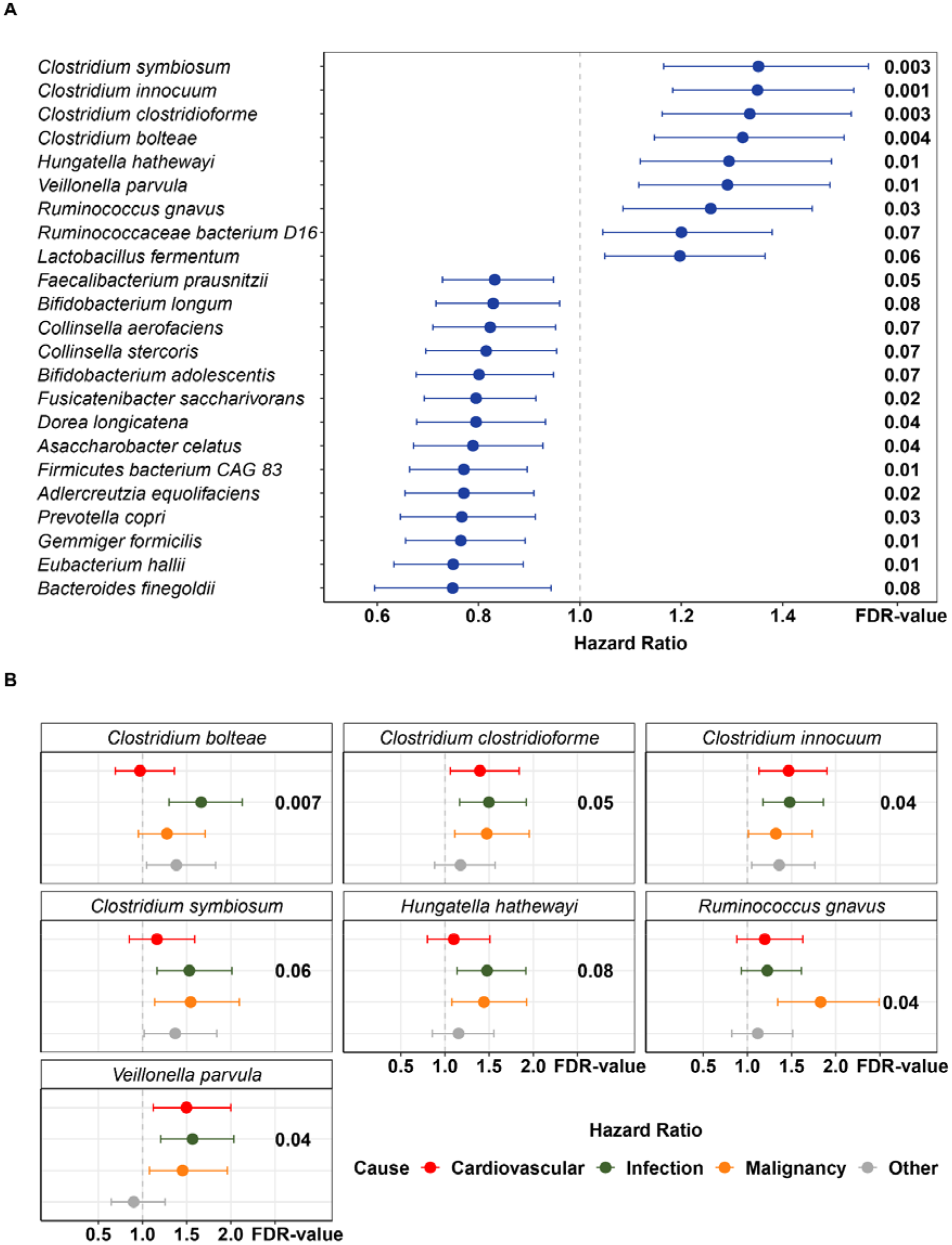
**(A)** Forest plot depicting all-cause mortality associated species from Cox-regression analysis. Hazard-ratio and 95% confidence interval and the FDR-corrected value are shown. **(B)** Forest plot depicting cause-specific mortality associated species from Cox-regression analysis. Hazard-ratio and 95% confidence interval and the FDR-corrected value are shown.

We took the analyses one step further by categorizing species based on whether their CLR-transformed abundance in each transplantation recipient were outside of its ‘normal’ range in the general population (higher [>75% quantile] or lower [<25% quantile]) and whether this binary classification associated more strongly with mortality. This measure is relatively similar to our dysbiotic indicator ‘distance to the average of the general population’ but instead gives an indication of dysbiosis on the species as opposed to the microbial community level. When we analyzed all SOTR, we found many of the same associations (11/16 and 22/23 species at an FDR<0.05 and FDR<0.1, respectively) but with stronger hazard ratios (at an FDR<0.05; **Figure 4**). For example, the four *Clostridium* species (*C. innocuum, C. clostridioforme, C. symbiosum* and *C. bolteae*) that were positively associated with all-cause mortality in the previous analysis exhibited up to 1.8 times higher hazard ratios in this analysis (range of increase: min=1.3, median=1.35, max=1.8; **Figure 4**).

**Figure 4.**
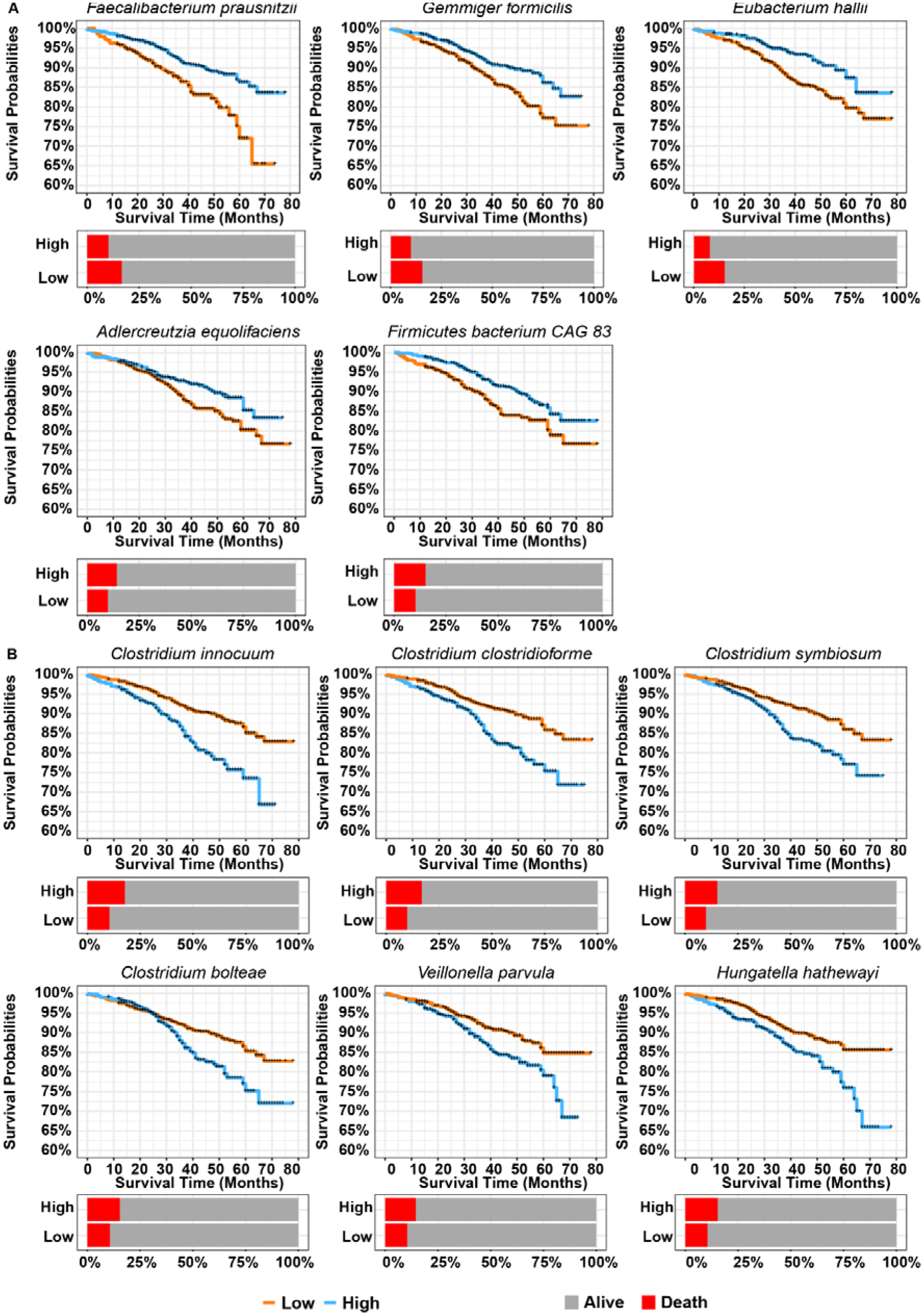
Kaplan-Meier curves showing the mortality probabilities of recipients compared with the general population. Bar plots depict the percent of deceased patients in the low and high group, respectively. **(A)** Decreased mortality risk; i.e., if the relative abundance of the bacteria of a SOTR corresponds to the highest quantile of the general population there was a significantly lower risk for mortality. **(B)** Increased mortality risk; i.e., if the relative abundance of the bacteria of a SOTR corresponds to the highest quantile of the general population there was a significantly higher risk for mortality.

### Gut microbiome as predictive biomarker of mortality

Finally, to identify a predictive biomarker we used a machine learning algorithm (CoDaCoRe). This algorithm has been developed to identify the log-ratio most predictive of an outcome, in this case death (dead/alive at the time of censoring) in all SOTR (**see Methods**). We first split our data into a training and testing set (80/20) with a proportional number of events in each set. This algorithm identified a log-ratio consisting of 19 species (AUC=0.68; **Figure 5A and 5B**) that had a classification accuracy of 88% in the test set. In this log-ratio, the numerator – or species whose joint abundance is predictive of death consisted of eight species (*Bacteroides eggerthii, B. fragilis, H. hathewayi, C. bolteae, C. clostridioforme, C. symbiosum, Ruminococcaceae bacterium D16* and *Escherichia coli*; **Figure 5A; Supplementary Table 9**), and the denominator – or species whose joint abundance is predictive of mortality consisted of 11 species (*B. adolescentis, B. longum, Adlercreutzia equolifaciens, A. celatus, P. copri. E. hallii, Anaerostipes hadrus, Coprococcus comes, D. longicatena, Fusicatenibacter saccharivorans* and *G. formicilis*; **Figure 5A; Supplementary Table 9**). All of these species were individually identified to be associated with mortality in our previous analyses (FDR<0.10; **Figure 3A**; **Supplementary Table 5**). Finally, we tested whether the identified log-ratio also could predict mortality in a multivariable Cox regression including age, sex, BMI and years since transplantation. We found that this log-ratio (i.e., harboring more of the numerator species and less of the denominator species) is indeed associated with an elevated mortality risk (HR=1.74,95% CI=1.48-2.04, P=1.60 x 10^−11^, **Figure 5C**). When we categorized individual transplant recipients based on whether they harbored lower or higher than the median value of this log-ratio across all SOTR, we found that the hazard ratio increased by almost a factor of 1.4 (HR=2.40, 95% CI=1.71-3.35, P=3.40 x 10^−7^, **Figure 5D**).

**Figure 5.**
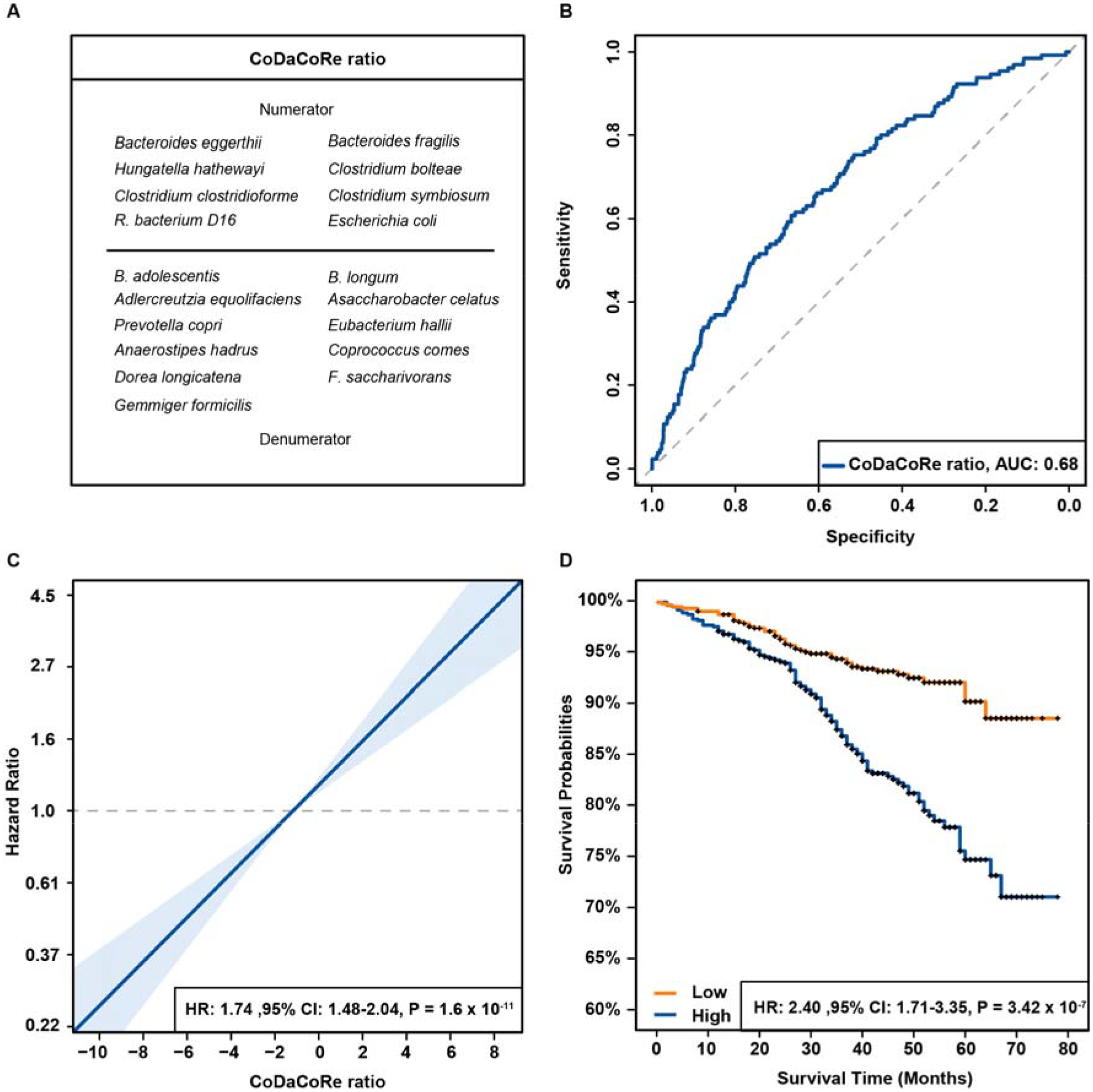
Machine learning algorithm identifies a log-ratio predictive of mortality. **(A)** Species included in the identified CoDaCoRe ratio. **(B)** AUC-ROC curve demonstrating discriminative power of the most predictive log-ratio identified by CoDaCoRe in the training set. **(C)** Blue line indicates the estimated hazard ratio compared to the log-ratio values with light blue area representing the 95% confidence interval (CI) of the hazard ratio. **(D)** Kaplan-Meier curves for recipients harboring lower (orange) and higher (blue) the median log-ratio value across all SOTR. R.: Ruminococcaceae, B.: Bifidobacterium, F.: Fusicatenibacter.

## Discussion

In the current study, we report an in-depth analysis of the gut microbiome in relation with both all-cause and cause-specific mortality in a population of SOTR from the TransplantLines cohort and biobank study.^13^ We observed gut microbial signatures associated with both all-cause and cause-specific mortality, especially death from infection. The distance of the gut microbiome to general population controls, resistome and virulence factor richness were associated with a higher mortality risk. We found a consistent mortality related gut microbial signal consisting of previously disease-associated species. Interestingly, we discovered that if the abundance of a species among SOTR is outside what is considered the ‘normal’ or ‘healthy’ range in the general population, it also predicts mortality. Overall, our results show that gut dysbiosis related gut microbial signatures are associated with mortality across different SOTR.

Diversity analysis was partly consistent with previously reported results. Patients with a lower Shannon diversity index had 29% higher risk of malignancy related mortality. However, we did not observe a significant relationship between the Shannon diversity index and all-cause mortality in all the pooled SOTR analysis or stratified by organ-type. Thus, we are unable to confirm previous reported associations between the Shannon diversity index and mortality in HSCT-recipients and liver transplant recipients.^10,11^ Similar to the gut microbiome mortality analysis in the general population, we observed a significant relationship between all-cause and cause-specific mortality and the PCA signature.^8^ SOTR that were one standard deviation higher than average in PC1 had a 32% higher mortality risk while SOTR lower in PC3 had a 20% lower mortality risk. We observed a consistent gut microbial signal in all of our mortality analyses, i.e., the PCA mortality analysis, the gut dysbiosis indicator analysis, and the per-species and machine learning mortality analysis.

Many of the species that were associated with mortality in our study, have previously been associated with disease in the general population.^5^ Specifically, we found that abundances of several clostridium species, including *C. clostridioforme, C. symbiosum, C. bolteae*, and *V. parvula* and *R. gnavus* were significantly associated with higher mortality in SOTR and with multiple diseases in the general population. In contrast, *P. copri, D. longicatena and F. prausnitzii* were associated with lower mortality in SOTR and with general health in the general population.^5^ Furthermore, we observed that the extent of dissimilarity of the gut microbiome compared with the general population is associated with all-cause-, infection related-, cardiovascular related- and malignancy related mortality. This relationship was consistent with individual bacterial species in our study, but when compared to in the FINRISK study we were only able to confirm the observation of a lower mortality risk for SOTR with a higher abundance of *Faecalibacterium prausnitzii*.^8^ However, in the FINRISK study Cox-regression analysis was performed on the *genus* level and SHOGUN was used for taxonomical classification while we performed our analysis on the *species* level and used MetaPhlAn for taxonomic profiling, potentially limiting the comparability between the two studies.^14^ Thus, further research using diverse populations and standardized methodology is needed to test whether our findings generalize to broader populations.

We observed a lower abundance of four butyrate producing bacteria is linked to increased mortality; *Gemmiger formicilis, Firmicutes bacterium CAG 83, Eubacterium hallii* and *Faecalibacterium prausnitzii*.^15–18^ Butyrate is a short-chain fatty acid (SCFA) with a broad range of functionality including; microbiome modulation, anti-inflammatory activity, anti-obesity effect and antioxidant functions.^19^ It was previously observed that KTR and LTR have a lower abundance of butyrate producing bacteria compared with controls and that a lower abundance of butyrate producing bacteria is associated with a lower health related quality of life in KTR. ^11,20–22^ We now find a higher mortality risk for SOTR with a lower abundance of butyrate producing bacteria. These results suggest that reduced butyrate levels could potentially have a direct role in mortality for SOTR. A lower abundance of butyrate producing bacteria was associated with increased occurrence of graft-vs-host disease and transplantation related mortality in HSCT recipients.^23^ While measuring fecal SCFA was outside of the scope of the current study, future studies should evaluate fecal SCFA in relation with mortality. Our results warrant further studies into the role of butyrate producing bacteria and mortality in SOTR. The use of butyrate producing probiotics might offer a promising way to improve outcomes of solid organ transplantation.^24^

Strengths of the current study include a large sample size of the population of SOTR and the availability of a large control group from the Dutch population. With this dataset we were able to pinpoint a gut microbiome - mortality related signal in a group of SOTR with a high prevalence of dysbiosis. A limitation of the current study is that samples were not obtained at uniform time points after transplantation due to the cross-sectional nature of the cohort. Furthermore, we report results from an observational study which limits us to identify any causal relationships. It is possible that reverting dysbiosis will improve survival after transplantation, but it is also possible that the gut microbial signature that we observe is the effect of poor overall health and that the effect is not causal.

This study highlights a dysbiosis gut microbial - mortality signal in a population of SOTR with a high prevalence of dysbiosis. These findings are of interest for the transplant community as well as our general understanding of the relationship between the gut microbiome and health. Our results support emerging evidence showing that gut dysbiosis is predictive of long-term survival, indicating that gut-microbiome targeting therapies might improve patient outcomes although causal links should be identified first.

## Methods

### Study design

All SOTR cross-sectional gut microbiome data from the TransplantLines Biobank and Cohort study (Trial registration number NCT03272841) was included.^13^ The TransplantLines study has been previously described in detail and aimed to include all potential adult solid organ transplant recipients and kidney donors at the University Medical Center Groningen (UMCG), The Netherlands, starting from June 2015.^13^ We included 1337 fecal samples from SOTR. 8,208 subjects from the Dutch Microbiome Project were included to quantify the extent of dysbiosis and per species dysbiosis analysis.^5^ Fecal samples from TransplantLines and DMP were collected using the same procedures and processed with the same DNA extraction protocols (see below). All participants signed an informed consent form prior to sample collection. TransplantLines (METc 2014/077) and Lifelines (METc 2017/152) were approved by the local institutional ethics review board (IRB) from the UMCG. Both studies adhere to the UMCG Biobank Regulation and are in accordance with the World Medical Association (WMA) Declaration of Helsinki and the Declaration of Istanbul.

### Clinical data

In the TransplantLines study, every transplant recipient was asked to fill in questionnaires and blood, urine and fecal samples were collected. A detailed description, including details regarding the rationale of the study design, inclusion/exclusion criteria and randomization of the TransplantLines study is given by Eisenga *et al*..^13^ In the current study, the primary outcome was overall survival. Clinical records were assessed to verify if a participant was alive or deceased, using a censoring date of January 1^st^ 2022. If a patient was deceased, we assessed the cause of death and classified the cause of death into cardiovascular, infection, malignancy or other related mortality categories.

### Sample selection and gut microbiome data generation

#### Fecal sample collection and subsequent processing

Patients were asked to collect a fecal sample the day prior to the study visit. A FecesCatcher (TAG Hemi VOF, Zeijen, The Netherlands) was sent to the patients at home. Feces were collected and stored in appropriate tubes and frozen at home (at -18°C) immediately after collection. Frozen fecal samples were collected by UMCG personnel and stored at -80°C until DNA extraction.

#### DNA extraction

Microbial DNA was extracted using QIAamp Fast DNA Stool Mini Kit (Qiagen, Germany) according to the manufacturer’s instructions. The QIAcube (Qiagen, Germany) automated sample preparation system was used for this purpose. Library preparation was performed using NEBNext® Ultra™ DNA Library Prep Kit for Illumina for samples with total DNA amount lower than 200ng, as measured using Qubit 4 Fluorometer, while samples with DNA yield higher than 200ng were prepared using NEBNext® Ultra™ II DNA Library Prep Kit for Illumina®. Libraries were prepared according to the manufacturer’s instructions. Metagenomic shotgun sequencing was performed using Illumina HiSeq 2000 sequencing platform and generated approximately 8 Gb of 150 bp paired-end reads per sample (mean 7.9 gb, st.dev 1.2 gb). Library preparation and sequencing were performed at Novogene and MGI.

#### Metagenomic data processing

Illumina adapters and low-quality reads (Phred score <30) were filtered out using KneadData (v0.5.1)^25^. Then Bowtie2 (v2.3.4.1)^26^ was used to remove reads aligned to the human genome (hg19). The quality of the reads was examined using FastQC toolkit (v0.11.7). Taxonomy alignment was done by MetaPhlAn3 (v3.7.2)^26,27^ with the database of marker genes mpa_v20_m200. Metacyc pathways were profiled by HUMAnN2 (v0.11.1)^28^. Bacterial virulence factors and antibiotic resistance genes were identified using shortBRED [shortbred_identify.py (v0.9.5) (51) and shortbred_quantify.py tool (v0.9.5)] against virulence factors of pathogenic bacteria (VFDB) database (http://www.mgc.ac.cn/VFs/main.htm) and comprehensive antibiotic resistance database (CARD) (https://card.mcmaster.ca/) separately. Samples were further excluded in case of an eukaryotic or viral abundance >25% of total microbiome content or a total read depth <10 million. In total, we identified 1132 taxa (17 phyla, 27 class, 52 order, 98 family, 231 genera and 705 species). After filtering for a prevalence of 10% and relative abundance threshold of 0.01%, 141 species were left. Hereafter, total-sum normalization was applied. Analyses were performed using locally installed tools and databases on CentOS (release 6.9) on the high-performance computing infrastructure available at UMCG and University of Groningen (RUG). An example of scripts used for microbiome processing is available at https://github.com/GRONINGEN-MICROBIOME-CENTRE/TransplantLines.

#### Statistical analysis

Centered log-ratio normalization was used due to the compositional nature of the metagenomic sequencing data^29^. PCA was performed using Euclidean distance between clr-transformed abundances (Aitchison distance^30^) of bacterial species. The Shannon diversity index was calculated using the *vegan*^31^ package in R. Cox proportional hazard models using the R packages survival and rms was used including age, sex, BMI and years since transplantation to analyze the association between diversity metrics and mortality. We used the *cox_wrapper* function from Salosensaari *et al*. to analyze the relationship between the gut microbiome and mortality per species.^8^ To further analyze the relationship between dysbiosis and the gut microbiome we used gut microbiome data from the general population and reclassified species abundance for SOTR according to quantiles of the general population. Cox-regression analysis was performed on this newly classified data.^5^ To assess which ratio of bacteria best predicted mortality in SOTR we applied CoDaCore with mortality status as the dependent variable using balances as the log-ratio type, a lambda of 0 and a maximum for base learners of 1.^32^ The most predictive ratio was than calculated per SOTR and Cox-regression analysis was performed on this ratio. False discovery rate was applied as a correction for multiple testing in all analysis.^33^

## Data availability

The raw microbiome sequencing data and basic phenotypes used in this study are available at the European Genome-Phenome Archive under accession numbers EGAD00001008907 (https://ega-archive.org/datasets/EGAD00001008907), EGAS00001006257 (https://ega-archive.org/studies/EGAS00001006257) and EGAS00001006258 (https://ega-archive.org/studies/EGAS00001006258). Due to patient confidentiality, the clinical datasets associated with the metagenomic datasets are available upon request to the University Medical Centre Groningen. Access to this clinical dataset requires a minimal access procedure consisting of a request per email for a data access form. A response will be provided within two working weeks. This access procedure is to ensure that the data are being requested for research/scientific purposes only and thus complies with the informed consent signed by TransplantLines participants, which specifies that the collected data will not be used for commercial purposes.

## Acknowledgements

We would like to thank all participants from the TransplantLines- and Lifelines cohort and biobank study. We would like to thank the Center for Information Technology of the University of Groningen (RUG) for support and for providing access to the Peregrine high-performance computing cluster and the Genomic Coordination Center (UMCG and RUG) for support and for providing access to Calculon and Boxy high-performance computing clusters. The TransplantLines Biobank and Cohort study received funding from Astellas BV (TransplantLines Biobank and Cohort study) and Chiesi Pharmaceuticals BV (PA-SP/PRJ-2020-9136) and was co-financed by the Dutch Ministry of Economic Affairs and Climate Policy by means of the PPP-allowance made available by the Top Sector Life Sciences & Health to stimulate public-private partnerships. Sequencing of the kidney part of the TransplantLines cohort was funded by a grant from the Dutch NWO/TTW/DSM partnership program Animal Nutrition and Health (project number 14939) to S.J.L.B. R.K.W. is supported by the Seerave Foundation, the Netherlands Organization for Scientific Research (NWO), and the EU Horizon Europe Program grant miGut-Health: personalized blueprint of intestinal health (101095470).

