## Supplementary Materials for "Multiple indicators of gut dysbiosis predict all-cause and cause-specific mortality in solid organ transplant recipients"

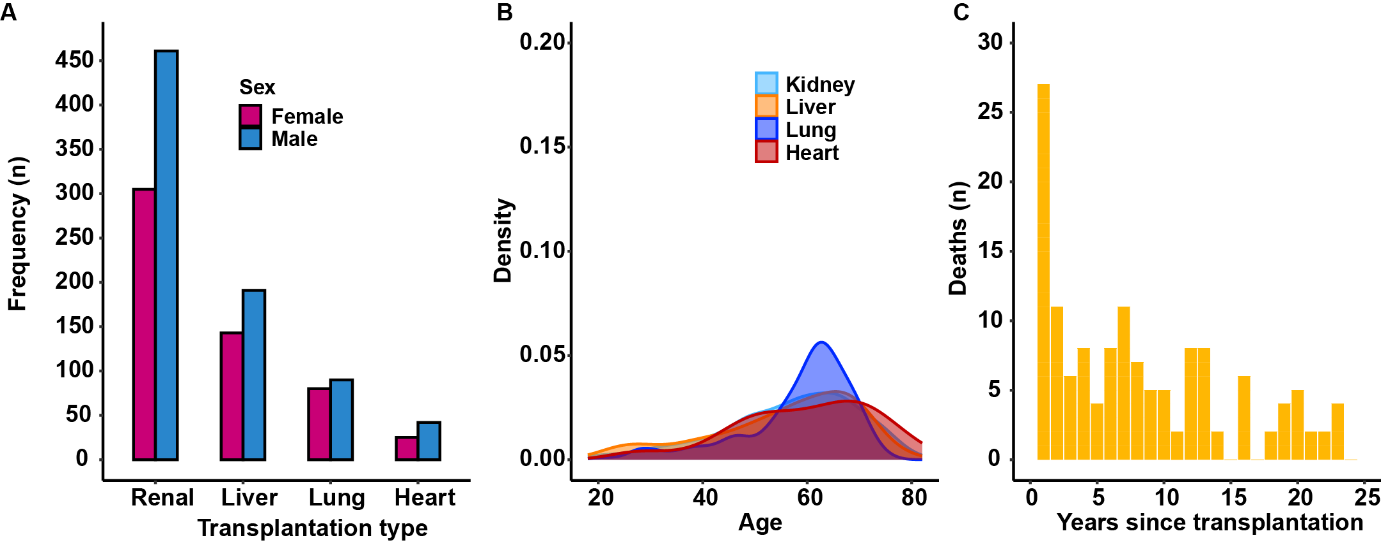

**Supplementary Figure 1** **(A)** Baseline characteristics of all SOTR including sex, **(B)** age and **(C)** the number of deaths.

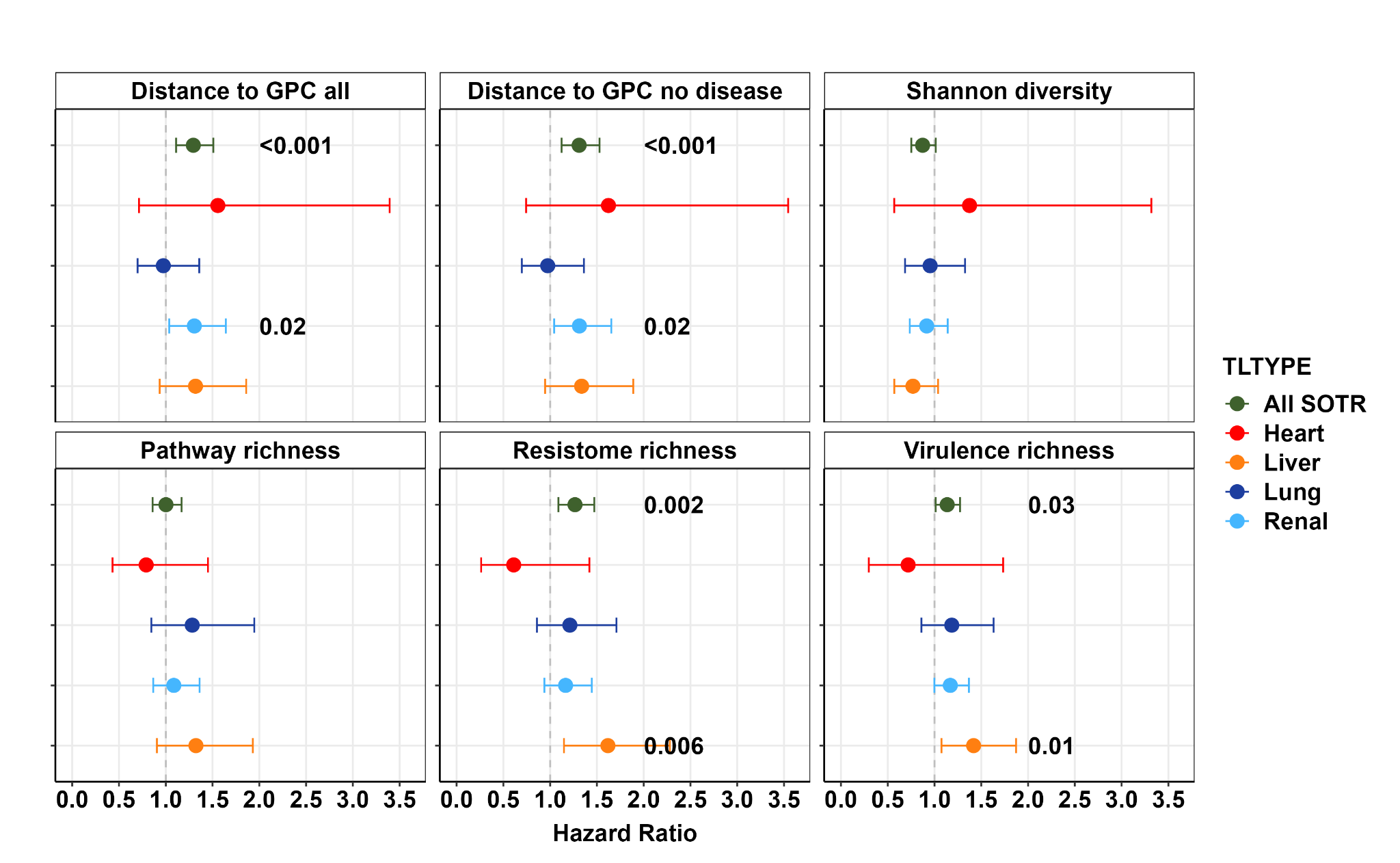

**Supplementary Figure 2** Alpha-diversity metrics stratified per transplantation type.
